# Gender and ethnicity intersect to reduce participation at a large European Hybrid HIV Conference

**DOI:** 10.1101/2023.02.01.23285329

**Authors:** A. Howe, YI. Wan, Y Gilleece, K Aebi-Popp, R Dhairyawan, S Bhagani, S. Paparini, C. Orkin

**Affiliations:** SHARE Collaborative, Wolfson Institute of Population Health, Queen Mary University of London, London UK; William Harvey Research Institute, Queen Mary University of London, London, UK, EC1M 6BQ; Acute Critical Care Research Unit, Royal London Hospital, Barts Health NHS Trust, London, UK, E1 1FR; Department of Infectious Diseases, Bern University Hospital, University of Bern, Switzerland; SHARE Collaborative, Blizard Institute, Queen Mary University of London, London UK; Medical Women’s Federation, BMA House, London, UK; Barts Health NHS Trust, London, UK; Royal Free London NHS Trust, London, UK; University College London, London, UK; European AIDS Clinical Society (EACS), Brussels

**Keywords:** Gender, ethnicity, inequalities, intersectionality, medical conferences, inequity

## Abstract

**Introduction:** The negative effect of female gender identity on participation at face-to-face academic conferences for delegates, speakers, chairs and panellists has previously been reported. Little is known about how ethnicity may affect conference participation, or about how gender and ethnicity intersect. To our knowledge, this is the first study describing conference participation by both ethnicity and gender in panellists and delegates, and the first to describe this within a hybrid conference setting.

**Methods:** We collaborated with the European AIDS Clinical Society (EACS), the organisers of the 18th European AIDS Conference, a large, 3223 delegate, hybrid conference held online and in London, over four days in October 2021. During the conference, we observed the number and type of questions asked at 12 of 69 sessions and described characteristics of the panel composition by ethnicity, gender and seniority. A post-conference survey of conference attendees collated demographic information, number of questions asked during the conference and the reasons for not asking questions.

**Results:** Men asked the most questions and were more likely to ask multiple questions in the observed sessions (61.5%). People from White ethnic groups asked >95% of the questions in the observed sessions. The fewest questions were asked in the sessions with the least diverse panels in terms of both ethnicity and gender. Barriers to asking questions differed between genders and ethnicities.

**Conclusions:** Improvement in access and participation at medical conferences is needed. Our study aims to raise awareness and provide evidence to help improve equality, diversity and inclusion in the professional medical conference setting and support equitable dissemination and sharing of knowledge. Intersections of gender and ethnicity shape inequality and need to be examined in combination. Further work is needed to evaluate the role of other social identities. We recommend future work takes such intersectionality into account and that conference organisers strive for diversity on panels to improve education and engagement of delegates.

## Introduction

Participating in academic conferences is integral to academic life and offers opportunities for education, knowledge dissemination, shared learning, visibility, collaboration and networking.[1] In a recent survey of the UK workforce, women represented only 19% of professors, 37% of senior academics and 44% of lecturers and only 0.7% of all UK professors were both Black and female, evidencing how gender and race can intersect to amplify inequity.[2, 3] Barriers to attendance of conferences may exacerbate inequalities, and widen education and professional attainment gaps.[4] Since the pandemic, international conferences have been delivered both online, face-to-face and as ‘hybrid’ (online and in-person) meetings. This has expanded opportunities to attend for those who may otherwise have been excluded by barriers such as caring duties or visa requirements. [5, 6]

In terms of the effects of gender, a greater prominence of male versus female academic speakers and chairs at medical conferences has been reported globally and across many specialities.[7-9] With respect to participation at conferences, assessing questions asked is a helpful proxy. One study showed that when men ask the first question, women ask proportionately fewer subsequent questions and ‘internal factors’ such as ‘*not being able to work up the nerve*’ were cited by women as barriers to asking questions.[10] Two studies at science conferences showed that men asked 80% more questions than their female colleagues even in a gender-balanced room.[9, 11] Within the field of medicine, Salem et al found that, in a gender-balanced room, women asked only 24% of questions at a national endocrinology conference.[12] Although the authors described the effects of seniority they did not evaluate the possible effects of ethnicity on participation.

Addressing the effects of ethnicity, a study by Bhayankaram and Bhayankaram evaluated ethnicity of panels and speakers at medical conferences.[13] They found that across twenty UK-based conferences, only 10% achieved an equal balance of speakers from White ethnic groups vs those from any other ethnic groups and at two in twenty, all the speakers were from White ethnic groups. [13] The hashtag ‘manel’ (all male panel) and ‘Whanel’ (all White panel) movements came into being, to call out such instances.[14, 15]

Wu et al. demonstrated that attendance at online meetings was higher than at face-to-face meetings with a more diverse representation of Black and Hispanic delegates from low and middle income countries.[16] However, the authors did not assess participation.

## Methods

We used a mixed-methods approach consisting of a narrative literature review of current evidence, a quantitative descriptive analysis of observed panel composition and audience member participation, and a post-conference survey of EACS 2021 participants. The research was co-created by researchers at Queen Mary University, the President and senior Officers of the European AIDS Clinical Society, and members of the Medical Women’s Federation.

### Literature Review

We carried out a narrative review of the available literature from a PubMed search, using the terms: “hybrid”, “medical”, “science”, “conference”, “meeting”, “congress”, “workshop” and included all results up to September 2022. Papers were included that analysed the hybrid format of a conference. Papers were excluded if they were related to the logistical/technical set up of a hybrid conference or about hybrid medical education. Identified papers were then analysed for any information about participation, gender, ethnicity, diversity, or inclusion in a hybrid setting.

### Observed Conference Participation

The 18^th^ European AIDS Conference was an international HIV conference, held in London, UK over four days in October 2021 with 3223 delegates and 69 sessions. With the collaboration of conference organisers, a researcher (AH) was facilitated to observe a total of 12 sessions (both hybrid and online). We chose a range of session types to include symposia, research abstract presentations, and parallel sessions. All observed sessions included time for questions and discussion, providing opportunity to assess participation. For session observations, a pre-structured observation tool (see Supplement S1) was used to gather data on panel and audience characteristics such as observed gender (we summarise here as male, female, unknown/other), observed ethnicity (we summarise into five broad ethnicity categories White, Black, Asian, Mixed and Other ethnic group based on the England and Wales 2021 Census classification),[17] country of work (UK, Europe, outside the European Union (EU), unknown/other) and level of career seniority. Information on observed gender and ethnicity were determined in three ways; reviewing the conference programme, online searches (Google) and direct observation. Limitations of this approach are discussed further on in the paper. Language of the question was classified into one of four themes; neutral, polite, shows under-confidence, shows over-confidence. The variables collected, codes, corresponding language and examples are given in Supplement Table S2. Descriptive statistics are presented as n (%). All data were collected Microsoft Excel (2021) and analyses were performed using R version 4.0.2 (R Core Team 2020).

### Post-Conference Survey

EACS secretariat staff emailed all delegates one month after the conference inviting them to complete an anonymous online survey about audience engagement with hybrid meetings. The survey collected data on age, self-reported gender, self-identified ethnicity, English proficiency, seniority, and preferences for on-site or virtual or hybrid meetings. We asked participants to self-identify by choosing from five options describing ethnicity derived from the England and Wales Census: [White, Black or Black British, Caribbean or African, Asian or Asian British, Mixed or Multiple ethnic groups, and or Other ethnic group].[17] For the sake of clarity in reporting, we once again use the broad ethnicity groupings Black, White, Asian, Mixed or Other. The survey was open for 28 days and collected data on number of questions asked and reasons for not asking questions. Survey questions and conduct are described in the supplementary material (Supplement S3). The QMUL Jisc online survey software was used.

## Results

### Literature Review

Of 99 records identified and screened, 25 papers were included in full text analysis. Of these, only seven met the inclusion criteria (See supplement S4, S5). The vast majority (6/7, 86%) of these papers were just simple surveys of satisfaction of the hybrid format. Only one of them gave a breakdown of gender of the participants but none analysed the effects of the hybrid format on the ethnicity or gender of the speakers or participants (Supplement S6).

### Conference Participation

Of 3223 attendees, 154 (4.8%) were faculty (speakers, chairs and panellists), 2,294 (71.2%) attended online and 929 (28.8%) on-site. The majority of delegates were from the UK and EU (67%), 12% from North America, 11% from Russia, 8% from South America and the remainder (1.2%) were from Africa and Australasia. The majority (89%) of the on-site delegates were from the EU and UK. Delegate numbers in 2019 were similar to 2021, 3145 vs 3223 respectively. We observed 12 out of a total of 69 sessions. Of these five were hybrid and seven were online only. We chose these at random from all the sessions that included question and answer sessions. Characteristics and composition of panel members by gender and ethnicity are shown in Table 1 and in Figure 1 respectively. There were 82 panel members, of whom 53.7% were female. Broken down by observed ethnicities, the researcher classified 72 (87.8%) as White, four (4.9%) as Black, and six (7.3%) as Asian. The majority (94%) of panellists came from the EU and the UK. The seniority of panellists is described Table 1.

**Table 1.** Characteristics of panel members by gender and observed ethnicity. Data presented as n (%).

| PANEL MEMBER | FEMALE | MALE | PANEL MEMBER | WHITE | BLACK | ASIAN |
| --- | --- | --- | --- | --- | --- | --- |
| n | 44 | 38 | n | 72 | 4 | 6 |
| *Session type online* | 29 (65.9) | 27 (71.1) | *Session type online* | 48 (66.7) | 3 (75.0) | 5 (83.3) |
| *Access in-person* | 12 (27.3) | 7 (18.4) | *Access in-person* | 17 (23.6) | 1 (25.0) | 1 (16.7) |
| *Gender female* | - | - | *Gender female* | 37 (51.4) | 2 (50.0) | 5 (83.3) |
| *Ethnicity* |  |  | *Ethnicity* |  |  |  |
| White | 37 (84.1) | 35 (92.1) | White | - | - | - |
| Black | 2 (4.5) | 2 (5.3) | Black | - | - | - |
| Asian | 5 (11.4) | 1 (2.6) | Asian | - | - | - |
| *Country* |  |  | *Country* |  |  |  |
| UK | 19 (43.2) | 8 (21.1) | UK | 22 (30.6) | 2 (50.0) | 3 (50.0) |
| Europe | 20 (45.5) | 28 (73.7) | Europe | 45 (62.5) | 2 (50.0) | 1 (16.7) |
| Outside EU | 5 (11.4) | 2 (5.3) | Outside EU | 5 (6.9) | 0 (0.0) | 2 (33.3) |
| *Seniority* |  |  | *Seniority* |  |  |  |
| Professor | 12 (27.3) | 7 (18.4) | Professor | 19 (26.4) | 0 (0.0) | 0 (0.0) |
| Doctor/Senior academic | 19 (43.2) | 22 (57.9) | Doctor/Senior academic | 37 (51.4) | 2 (50.0) | 2 (33.3) |
| Scientist/AHP/PhD student | 12 (27.3) | 5 (13.2) | Scientist/AHP/PhD student | 12 (16.7) | 1 (25.0) | 4 (66.7) |
| Non-medical professional | 1 (2.3) | 2 (5.3) | Non-medical professional | 2 (2.8) | 1 (25.0) | 0 (0.0) |
| Unknown/other | 0 (0.0) | 2 (5.3) | Unknown/other | 2 (2.8) | 0 (0.0) | 0 (0.0) |

**Figure 1.**
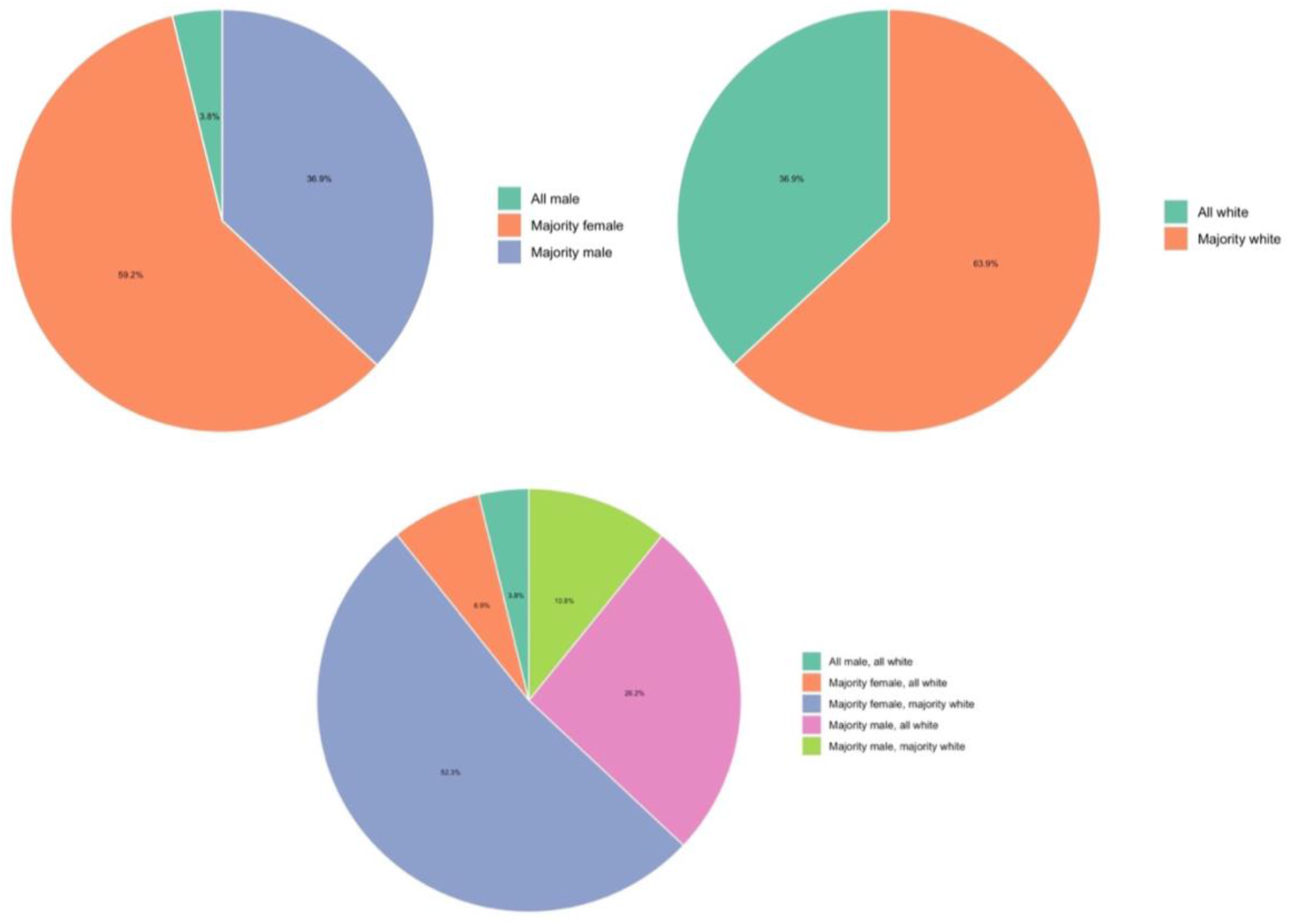
Session panel composition by gender and ethnicity. Total 12 sessions.

Across 12 sessions, 63 individuals (18 panel and 45 audience members) asked a total of 130 questions. A total of 44 (33.8%) questions were asked by the speaker/panel/chairs. Observed demographic characteristics of people who asked a question in any session are shown in Table 2. Men asked 57% of the questions (n=36) and those observed as White asked 88.9% (n=56) of the questions, where observed ethnicity was assigned. Twenty-seven people asked more than one question. Of those who asked more than one question (excluding one questioner with unknown characteristics), all were observed as White and 16 (61.5%) were male. Seven people asked five or more questions accounting for 30.8% of all questions asked, five were male and all were observed as White. During hybrid sessions, the chat function was used to ask between 44.4%- 100% of the questions.

**Table 2.** Characteristics of audience members who asked questions. Data presented as n (%). Excluding unknown gender (n=1) and unknown observed ethnicity (n=4).

| AUDIENCE MEMBER | FEMALE | MALE | AUDIENCE MEMBER | WHITE | BLACK | ASIAN |
| --- | --- | --- | --- | --- | --- | --- |
| n | 25 | 34 | n | 56 | 1 | 2 |
| *Session type online* | 13 (52.0) | 23 (67.6) | *Session type online* | 35 (63.5) | 0 (0.0) | 1 (50.5) |
| *Mode* |  |  | *Mode* |  |  |  |
| Microphone | 3 (12.0) | 2 (5.9) | Microphone | 5 (8.9) | 0 (0.0) | 0 (0.0) |
| Chat | 12 (48.0) | 24 (70.6) | Chat | 33 (58.9) | 1 (100.0) | 2 (100.0) |
| Chair/panel member | 10 (40.0) | 8 (23.5) | Chair/panel member | 18 (32.1) | 0 (0.0) | 0 (0.0) |
| *Gender female* | - | - | *Gender female* | 24 (42.9) | 1 (100.0) | 0 (0.0) |
| *Ethnicity* |  |  | *Ethnicity* |  |  |  |
| White | 24 (96.0) | 32 (94.1) | White | - | - | - |
| Black | 1 (4.0) | 0 (0.0) | Black | - | - | - |
| Asian | 0 (0.0) | 2 (5.9) | Asian | - | - | - |
| *Country* |  |  | *Country* |  |  |  |
| UK | 0 (0.0) | 1 (2.9) | UK | 1 (1.8) | 0 (0.0) | 0 (0.0) |
| Europe | 10 (40.0) | 12 (35.3) | Europe | 19 (33.9) | 1 (100.0) | 2 (100.0) |
| Outside EU | 14 (56.0) | 20 (58.8) | Outside EU | 34 (60.7) | 0 (0.0) | 0 (0.0) |
| Unknown | 1 (4.0) | 1 (2.9) | Unknown | 2 (3.6) | 0 (0.0) | 0 (0.0) |

| *Seniority* |  |  | *Seniority* |  |  |  |
| --- | --- | --- | --- | --- | --- | --- |
| Professor | 9 (36.0) | 5 (14.7) | Professor | 14 (25.0) | 0 (0.0) | 0 (0.0) |
| Doctor/Senior academic | 13 (52.0) | 20 (58.8) | Doctor/Senior academic | 30 (53.6) | 1 (100.0) | 2 (100.0) |
| Scientist/AHP/PhD student | 0 (0.0) | 3 (8.8) | Scientist/AHP/PhD student | 3 (5.4) | 0 (0.0) | 0 (0.0) |
| Non-medical professional | 1 (4.0) | 4 (11.8) | Non-medical professional | 5 (8.9) | 0 (0.0) | 0 (0.0) |
| Unknown/other | 2 (8.0) | 2 (5.9) | Unknown/other | 4 (7.1) | 0 (0.0) | 0 (0.0) |

The majority of the questions (n=84, 64.6%) were asked during the purely online sessions and by men (n=78, 60.4%, excluding 1 unknown). Delegates observed as White asked 98% (n=123) of the questions (excluding 4 unknowns). The relationship between the number of questions asked and the panel composition categorised by gender and observed ethnicity are shown in the supplement (S6-10) and Figure 2. Of the 12 sessions observed, seven of the panels were, by gender, majority (50% or more) female, four majority male, and one all male; and by observed ethnicity six were majority White and six all White. Mixed ethnicity panels comprised 10%-30% panel members from Black and Asian ethnicities. There was one all White, all-male panel (Figure 2).

**Figure 2.**
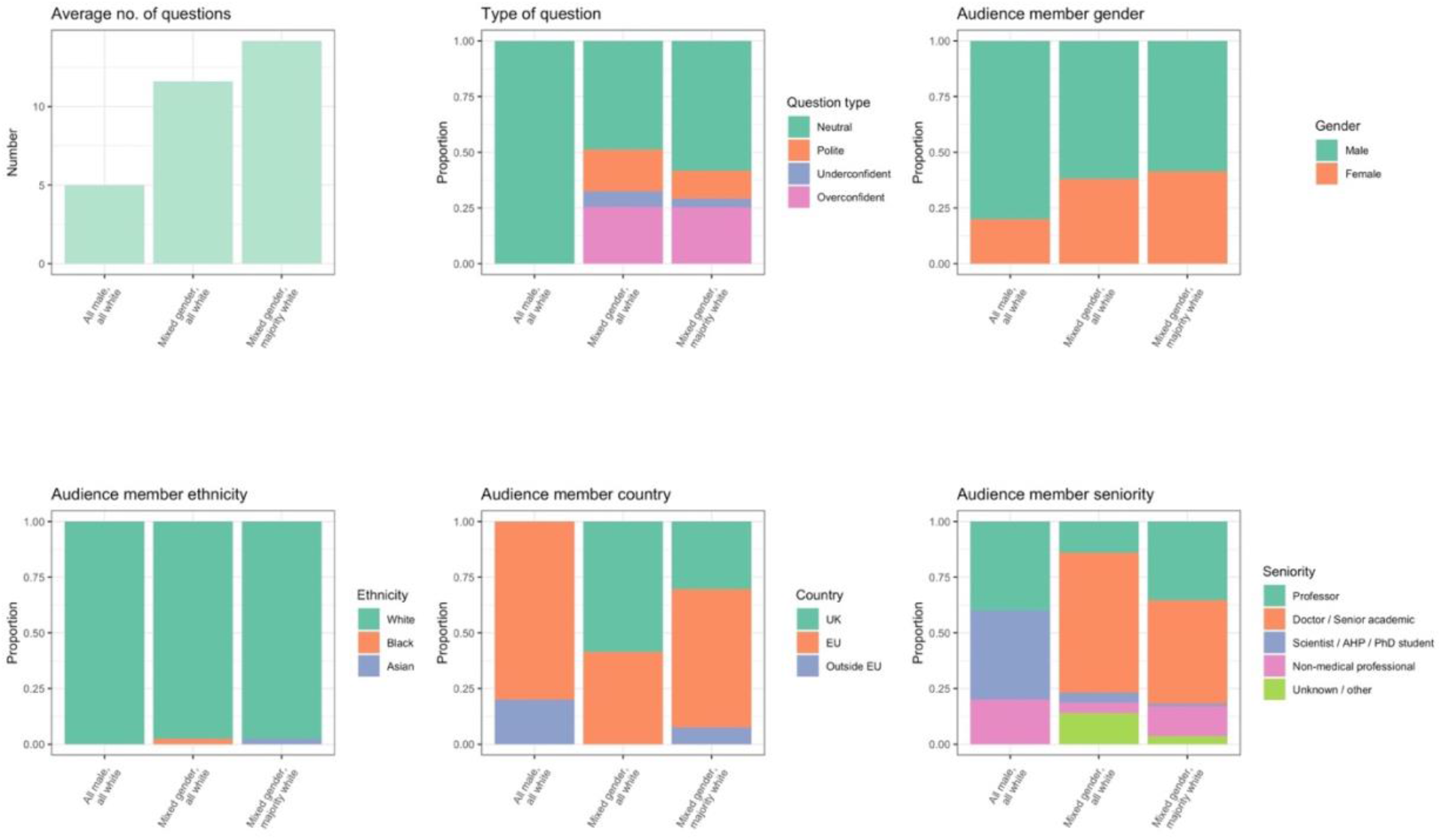
Relationship between panel composition by observed gender and observed ethnicity and number and type of questions asked by delegates. Total 12 sessions: 1 all male all White, 5 mixed gender all White, 6 mixed gender majority White.

With respect to the language used, four themes were identified; neutral, polite, shows under-confidence, shows over-confidence (see S2). Questions categorised as ‘over-confident’ were mainly asked by men (n=20, 64.5%), and by those observed as White (n=29, 93.5%). The majority of over-confident questions were asked using the chat function (n=19, 61.3%).

### Post Conference Survey

The survey completion rate was 17% (n=556/3223). Three-hundred and four (55.4%) identified as female. By ethnicity, 408 self-identified as White (74.7%), 36 (6.6%) as Asian, 21 (3.8%) as Black, 38 (7%) as mixed, and 43 (7.9%) identified as ‘other’. Almost 80% (n=419) of respondents were senior doctors or academics. Based on self-identification, three quarters (n=314) of the senior doctors or academics identified as White. Only seven of the senior respondents identified as Black- – they were all consultant grade, none were professors or senior academics. The remaining Black respondents were allied health professionals (n=7,1.3%) or community members (n=6, 1.1%).

White and male respondents had attended more prior conferences than other respondents (Table 3). Slightly more women preferred hybrid or online events versus men (32% vs 26%). Males and people who identify as Black showed preference for the microphone when asking a question (Table 3). By ethnicity, four people reported asking more than ten questions, all of whom self-identified as White.

**Table 3.** Questions answered in survey broken down by self-identified gender and ethnicity, as reported by the respondents of the online survey.

| Gender |  | Ethnicity |  |  |  |  |
| --- | --- | --- | --- | --- | --- | --- |
| Male | Female | White | Black | Asian | Mixed | Other |
| Have been to 21+ previous conferences |  |  |  |  |  |  |
| n=136, 60% | n=130, 42.8% | n=220, 54% | n=4, 19% | n=14, 39% | n=18, 47.4% | n=21, 48.8% |
| Preference for Hybrid or Online (vs in-person or no preference) |  |  |  |  |  |  |
| n=60, 26% | n=97, 32% | n=124, 30.4% | n=2, 9.5% | n=16, 44.4% | n=11, 29% | n=10, 23.3% |
| Preference for asking questions at the microphone (vs online) |  |  |  |  |  |  |
| n=38, 16.7% | n=24, 7.9% | n=51 (12.5%) | n=6 (28.6%) | n=3 (8.3%) | n=1 (2.6%) | n=6 (14%) |
| Asked 1 or more question at EACS 2021 |  |  |  |  |  |  |
| n=89, 39.2% | n=106, 34.9% | n=147, 36% | n=11, 52.4% | n=13, 36.1% | n=15, 39.5% | n=18, 41.9% |

The reasons for not asking questions differed by both gender and ethnicity, as shown in Table 4. More women reported being worried about asking a stupid question (n=63, 12.6% vs n=27, 8.1%), were more likely to feel shy or embarrassed (n=60, 12.1% vs n=24, 7.3%), or have a lack of confidence in their knowledge (n=36, 7.2% vs n=16, 4.8%) than men. More men, compared to women, cited not asking a question because they did not want to appear arrogant or critical, (n=20, 6% vs n= 8, 1.6%).

**Table 4.**
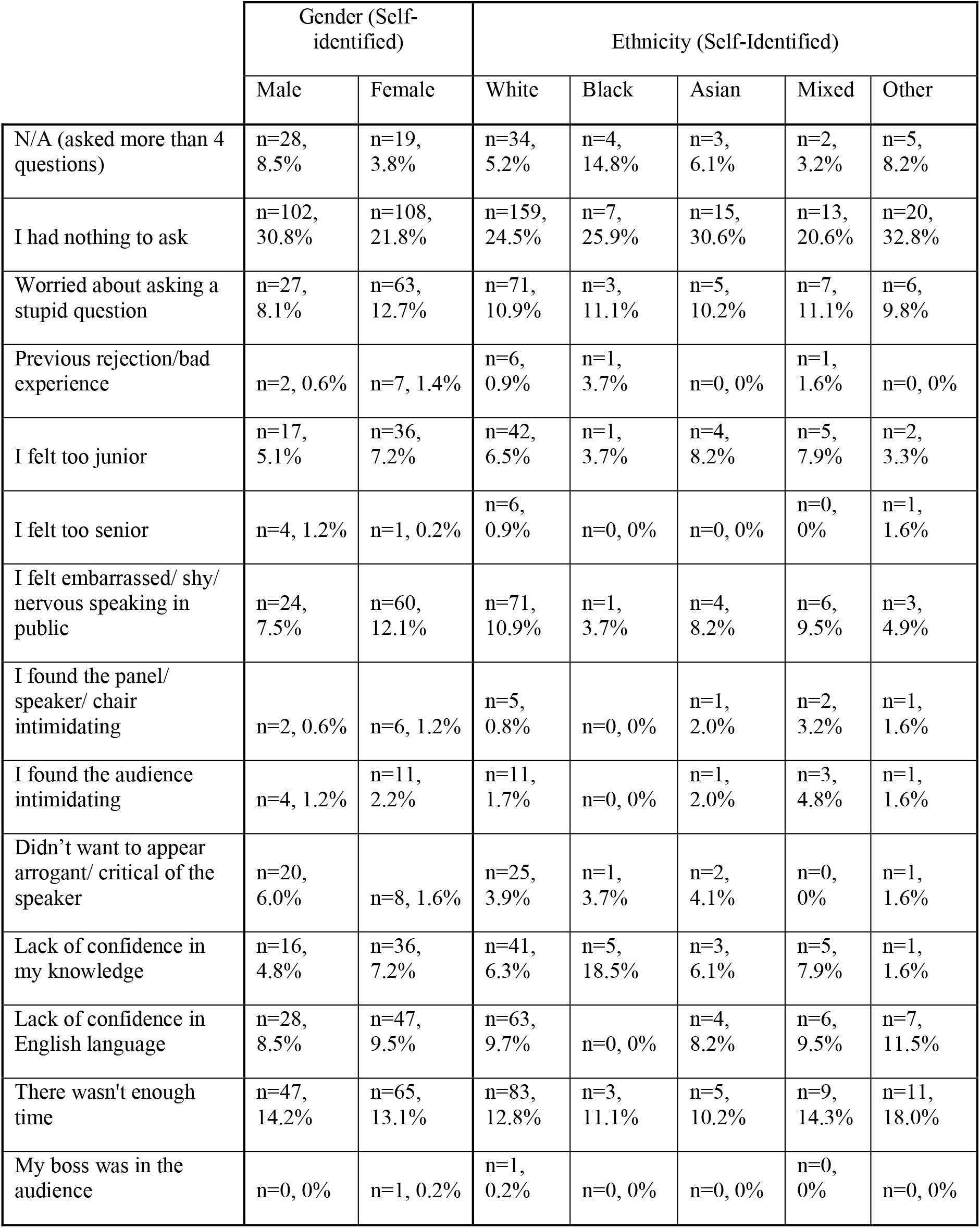
Barriers to asking questions at EACS 2021, broken down by self-identified gender and ethnicity, reported by respondents of the online survey. (Data removed for respondents who did not give gender or ethnicity)

|  | Gender (Self-identified) |  | Ethnicity (Self-Identified) |  |  |  |  |
| --- | --- | --- | --- | --- | --- | --- | --- |
|  | Male | Female | White | Black | Asian | Mixed | Other |
| N/A (asked more than 4 questions) | n=28, 8.5% | n=19, 3.8% | n=34, 5.2% | n=4, 14.8% | n=3, 6.1% | n=2, 3.2% | n=5, 8.2% |
| I had nothing to ask | n=102, 30.8% | n=108, 21.8% | n=159, 24.5% | n=7, 25.9% | n=15, 30.6% | n=13, 20.6% | n=20, 32.8% |
| Worried about asking a stupid question | n=27, 8.1% | n=63, 12.7% | n=71, 10.9% | n=3, 11.1% | n=5, 10.2% | n=7, 11.1% | n=6, 9.8% |
| Previous rejection/bad experience | n=2, 0.6% | n=7, 1.4% | n=6, 0.9% | n=1, 3.7% | n=0, 0% | n=1, 1.6% | n=0, 0% |
| I felt too junior | n=17, 5.1% | n=36, 7.2% | n=42, 6.5% | n=1, 3.7% | n=4, 8.2% | n=5, 7.9% | n=2, 3.3% |
| I felt too senior | n=4, 1.2% | n=1, 0.2% | n=6, 0.9% | n=0, 0% | n=0, 0% | n=0, 0% | n=1, 1.6% |
| I felt embarrassed/ shy/ nervous speaking in public | n=24, 7.5% | n=60, 12.1% | n=71, 10.9% | n=1, 3.7% | n=4, 8.2% | n=6, 9.5% | n=3, 4.9% |
| I found the panel/ speaker/ chair intimidating | n=2, 0.6% | n=6, 1.2% | n=5, 0.8% | n=0, 0% | n=1, 2.0% | n=2, 3.2% | n=1, 1.6% |
| I found the audience intimidating | n=4, 1.2% | n=11, 2.2% | n=11, 1.7% | n=0, 0% | n=1, 2.0% | n=3, 4.8% | n=1, 1.6% |
| Didn't want to appear arrogant/ critical of the speaker | n=20, 6.0% | n=8, 1.6% | n=25, 3.9% | n=1, 3.7% | n=2, 4.1% | n=0, 0% | n=1, 1.6% |
| Lack of confidence in my knowledge | n=16, 4.8% | n=36, 7.2% | n=41, 6.3% | n=5, 18.5% | n=3, 6.1% | n=5, 7.9% | n=1, 1.6% |
| Lack of confidence in English language | n=28, 8.5% | n=47, 9.5% | n=63, 9.7% | n=0, 0% | n=4, 8.2% | n=6, 9.5% | n=7, 11.5% |
| There wasn't enough time | n=47, 14.2% | n=65, 13.1% | n=83, 12.8% | n=3, 11.1% | n=5, 10.2% | n=9, 14.3% | n=11, 18.0% |
| My boss was in the audience | n=0, 0% | n=1, 0.2% | n=1, 0.2% | n=0, 0% | n=0, 0% | n=0, 0% | n=0, 0% |

## Discussion

This study is the first to describe both gender and ethnicity in panel composition, and the first to explore intersections between gender and ethnicity on number and types of questions asked at a large international hybrid meeting. Our research has shown that there are differences in participation by both gender and ethnicity and that this is related to the composition of the panel.

Considering gender alone, our findings concurred with previous studies in which despite a gender-balanced room, women asked fewer questions than men.[11] This may be due to a variety of reasons such as women’s concerns about their authority being doubted due to experiences of testimonial injustice and stereotypes about their gender.[18, 19] Like Salem et al we found that a significant proportion (40%) of the questions asked by women were asked by chairs/panellists.[12] This may be that these women who have already been chosen to speak may feel confident that they viewed as credible sources of information. In the observed sessions, although women represented 53.7% of the panellists, questions from men still outnumbered questions from women (57.6% vs 42.4%). Concerning ethnicity, people who were observed as White asked 95% of the questions. Out of 59 questions asked by female delegates, only one question was asked by an ethnically diverse woman. We showed that a gender-balanced panel alone was not enough to reach gender parity in the number of questions asked.

The composition of the panel seemed to be an important influencer of both number of questions asked per session and of who asked the questions. Looking at the panel composition through an intersectional lens, the panels were roughly gender balanced but 87.8% of panellists were observed as being White. Six panels were observed to be majority White and six were entirely White (‘all-White’). We found that the fewest questions were asked by anyone in the all-White panels compared with the more ethnically diverse panels. Fewer questions were asked by women in the all-White panels regardless of the gender balance (Figure 2.). There was one all-White, all-male panel. During this all-White, all-male session, the fewest questions were asked (5 vs an average of >10) for the other sessions. Four of the five questions asked and answered were by the panel members themselves and the single online question was asked by a woman and not answered. In summary, when there is both gender and ethnic diversity on the panel more questions were asked and women participated more. More men and more respondents from White ethnic groups asked over-confident questions and over-confident questions were mostly asked using the chat.

In the survey, we asked the reasons underlying the choice not to ask questions and found differences by both self-identified gender and ethnicity. More women than men reported feeling ‘shy or embarrassed’, ‘concerned about asking a stupid question’ and ‘insecure about their level of knowledge’. In the survey, women and Black respondents reported ‘lack of confidence in my knowledge’ as barriers to asking questions more frequently than men and people from other ethnic groups. There were ten (1.8%) people who reported that a previous bad experience or rejection was a barrier to asking a question. Of these, seven (70%) were female.

Concerning the hybrid format and the relationship between increased access and participation, our survey suggests that the former does not necessarily imply the latter. The majority (89%) of onsite attendees were from UK or EU which may reflect the restrictions of the COVID-19 pandemic as well as wider inequities for attendees from low-and-middle income countries. Black survey respondents had been to the fewest conferences but had the highest preference for in-person conferences, reported a strong preference for asking questions, and had the highest preference for asking questions at the microphone. Also, no Black respondents said that they preferred not to ask questions. This contrasts with what we observed during the twelve observed sessions where only one Black person asked a question and suggests a gap between what people would like to do versus what they actually did. We found that women have been to fewer conferences, have a higher preference for hybrid or online format and have a lower preference for asking questions at the microphone, compared to men.

### Limitations of this Research

We recognise that the main researcher and observer (AH) is a White cis-woman working as an academic clinical fellow in the United Kingdom and this may have influenced her observations.

We did not have access to the total number of attendees at the meeting by gender and ethnicity (the conference organisers do not collect such data), nor did we have these figures for the sessions we observed, so we were unable to relate the number of questions asked to the proportions of attendees. However, our study has shown that regardless of the denominator, panel composition appears to be an important influencer on overall audience participation with reduced panel diversity leading to fewer questions and fewer numbers of participants overall. The determination of the ethnicity and gender and seniority of the speakers and audience members was based on the observation of the researcher (AH) and corroborated by google searches where the information was not available on the conference programme, which may have led to inaccuracies or misrepresentation. However, the patterns we observed during the sessions were corroborated by the survey findings where ethnicity and gender were self-identified by respondents.

We presented the data on ethnicity as five broad ethnic groupings. We are aware that such reduction does not allow for a fully comprehensive understanding of the definition and role of ethnicity. However, we chose this way of presenting our data in order to be able to clearly show intersections with gender and other categories in our analysis in intelligible ways.

We also do not know what portion of EACS members or HCPs working in HIV Medicine (and related fields) are from racially minoritised groups, and whether the proportion who participated in this conference is representative of this as no such data exist.

In order to perform thematic analysis on the language used by participants asking questions, tone and manner were taken into consideration and a single code based on the dominant overall style of the question asked was used. The interpretation of the language used was based on the researchers (AH) interpretation.

The survey was only available in English which may have limited participation and it was open for a limited period (28 days). We did not provide the option for participants to indicate that they couldn’t ask questions because they had watched the session on-demand after the event had happened which was an omission and may have skewed the numbers of those who did not ask questions especially in people from different time zones.

## Conclusions

Equity of access and participation at medical conferences is an important component of achieving parity of experience, education, and professional development for women and minoritised groups. Our aim is to provide the first intersectional approach to ethnicity and gender in a hybrid medical conference. Ethnicity needs to be considered alongside gender, as intersectionality is a defining feature of inequality and identity categories are best not considered in a vacuum. Based on our findings which suggest that gender-balanced and ethnically diverse panels fostered greater engagement, we recommend that conference organisers actively strive to go beyond gender and embrace true diversity and inclusivity. Future research and interventions should evaluate and consider other structural barriers such as ability and other social identities. We recommend future work takes intersectionality into account and that conference organisers strive for diversity of speakers and on panels as a first step to improve engagement. We further recommend that conferences collect and publish diversity data at their conferences and that panellists are specifically briefed to ensure that they select questions from a diverse group of delegates.

## Data Availability

All data produced in the present study are available upon reasonable request to the authors

## Authors’ contributions

AH: collection of data, literature review, author of post conference survey, contributor to analysis of data, main author of paper.

YW: Analysis of data, main contributor/reviewer of text

YG: collaborator from EACS, final edits

KAP: collaborator from EACS, final edits

RD: contribution to drafts and final edits SB: collaborator from EACS, final edits

SP: contributor to analysis of qualitative data and main contributor/reviewer of text

CO: overall lead, supervisor, and main contributor/reviewer of text

## Author information

CO: President of Medical Women’s Federation

SB: President of EACS

YG: Senior Officer EACS

KA-P: Senior Officer EACS

## Acknowledgements

nil

## Disclaimer

nil

## Additional files

Additional file 1: Gender and ethnicity intersect to reduce participation at a large European Hybrid HIV Conference: Supplementary Material

Word Document

Additional supplementary tables and figures relating to the research.

