## Supplementary Materials for "Gender and ethnicity intersect to reduce participation at a large European Hybrid HIV Conference"

Supplementary Material

**Supplement S1.** Pre-structured observation tool used to gather information during the observed sessions.

Session Title:
Session Type:

| | Online/ in person | Gender | Ethnicity | Country | Profession |
|---|---|---|---|---|---|
| Speakers | | | | | |
| Chairs | | | | | |

| Questions | Mode (chat/ microphone) | Gender | Ethnicity | Country | Job | Answered? Yes/ no |
|---|---|---|---|---|---|---|
| 1 | | | | | | |
| 2 | | | | | | |
| 3 | | | | | | |
| 4 | | | | | | |
| 5 | | | | | | |
| 6 | | | | | | |
| 7 | | | | | | |
| 8 | | | | | | |
| 9 | | | | | | |

**Supplement S2.** Themes, corresponding language and codes and examples of language used in the 12 observed sessions

| Theme: | Language | Code | Examples |
|---|---|---|---|
| Neutral | Neutral comment or follow up question | 1 | What is the criteria used…?<br>Is there any evidence for…?<br>Are there any studies that….? |
| Polite | Gives compliment / praise<br>Asks permission<br>Gives thanks to speaker / panel | 2 | Thanks for an excellent review/presentation/talk<br>Thank you for your work…<br>I think it's really interesting how you…. |
| Shows under-confidence | Shows hesitancy<br>Apologises<br>Self-deprecates | 3 | Asking for guidance<br>'Maybe I missed it..' 'I hope I haven't missed something..'<br>'I can't pronounce in English' / 'sorry my English is bad'<br>'I just'/'please can I'…Wanted to add / wanted to say |
| Shows over-confidence | Talks about own research/ clinical practice/interests<br>Makes suggestion<br>Direct / challenging / patronising<br>Makes more than 1 point / asks multiple follow up questions<br>Showing off (stating things they've done or read) | 4 | 'I'm the director of…'<br>'The work *we* are doing/ I am leading on shows…'<br>Talking for a long time and asking multiple questions<br>'I want to make 2/3/4 comments about….'<br>'We need to be more clever…'<br>'I would really press people to…. / 'Studies I've read show…' (boasting)<br>Making a challenging or negative suggestion on the presenters work<br>Maybe you should do…x/y/z (challenging or negative) |
| - | Content data missing | 0 | |

**Supplement S3.** Questions included in the post-conference survey

Q1. Age
- 18-25
- 26-30
- 31-34
- 35-40
- 41-50
- 51-60
- 60+

Q2. Gender
- Cis-Female (assigned female at birth and identify as female)
- Cis-Male (assigned male at birth and identify as male)
- Transfemale
- Transmale Non-binary / nonconforming
- Other
- Prefer not to say

Q3. Country of Residence
- White UK/White European/White Other
- Mixed or Multiple Ethnic Group
- Asian/ Asian British/ Asian Other
- Black African/ Black Caribbean/ Black British/Black Other
- Other Ethnic Group

Q4. Current role (tick all that apply)
- Allied Health Professional (nurse, physio, dietician, psychologist, health advisor)
- Medical Student
- Doctor in Training (non infectious disease trainee)
- Academic Trainee
- Infectious Disease/ GU Trainee
- Infectious Disease / GU Consultant
- Other Consultant (non ID)
- GP / Family Doctor / Primary Care Physician
- Senior Academic / Professor
- Social Scientist / Researcher
- Community Member

Q5. How would you describe your area of focus? (Tick all that apply)
- Clinical
- Epidemiology
- Public Health
- Social Science
- Basic Science
- Personal

Q6. Is English your first language?
- Yes
- No

If no, do you consider yourself fluent in English?

Q7. How many conferences have you attended in your career?
- 1-5
- 6-10
- 11-20
- 21+

Q8. Do you prefer to attend a conference virtually or in person?
- Virtually
- In person
- Hybrid
- Don't mind

Q9. How do you prefer to ask a question at a conference?
- In the app / online platform
- At the microphone / in person
- Combination of both
- I prefer not to ask questions

Q10. How many questions did you ask at EACS 2021 in total, via the microphone, the app or live chat? (approximately if you cannot recall)
- 0
- 1-3
- 4-6
- 7-9
- 10+

Q11. How did you ask your questions?
- N/A - I did not ask a question
- Online
- In person/ at the microphone
- Mixture of both

Q12. If you asked a question online, did you post using your name or anonymously?

Q13. How many of your questions were picked or answered in total?
- 1
- 2
- 3
- 4
- 5
- 6+
- N/A

Q14. If you did not ask a question, or asked less than 4, why? Select all that apply.

- Not applicable - I asked 4 or more questions
- I had nothing to ask
- I was worried about asking a stupid question
- I have had previous rejection or bad experience when asking a question
- I felt too junior
- I felt too senior
- I felt embarrassed / shy / nervous when public speaking
- I found the panel/speaker/chair intimidating
- I found the audience intimidating
- I didn't want to appear arrogant or critical of the speaker
- Lack of confidence in my knowledge
- Lack of confidence in the English language
- There wasn't enough time
- My boss was in the audience

Q15. Do you have any further comments about asking questions at conferences?

**Supplement S4.**

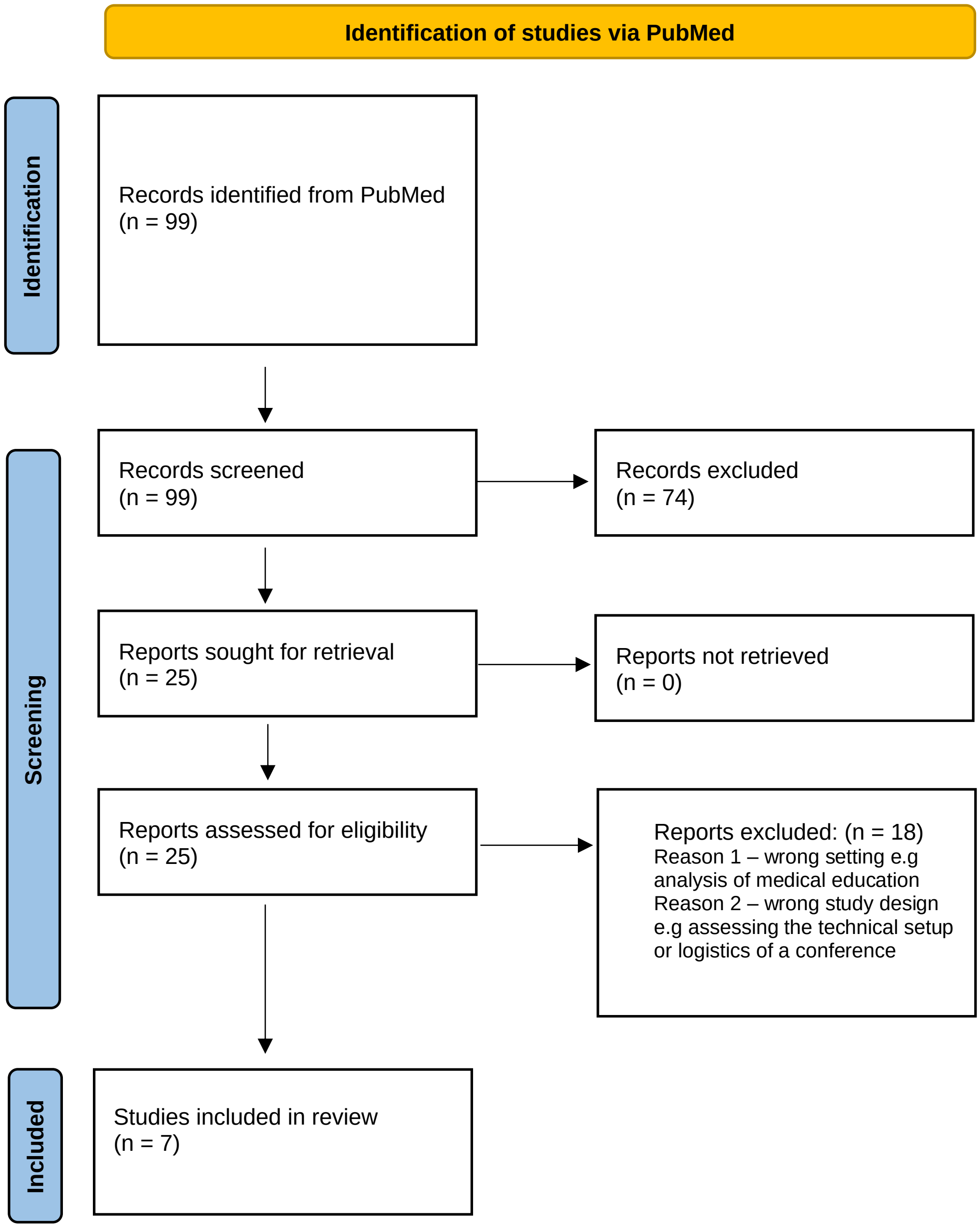

**Supplement S5.** The seven papers that met the inclusion criteria of the literature review.

| Title | Authors and Year | Specialty | Results | Comments | Gender or ethnicity mentioned? |
|---|---|---|---|---|---|
| Analysis of challenges faced, and the scientific content of a hybrid paediatric surgical conference arranged during theCOVID-19 pandemic | (Patel et al., 2021) | Paediatrics Surgery | 170 attendees. 90% in person attendance, 10% virtual. There were 107 (63%) male and 63 (37%) female participants. | Most participants were male but not statistically significant. No further analysis other than male vs female participants | yes (gender only) |
| ASNTR's Venture into a Hybrid Conference: Lessons Learned During the COVID-19 Pandemic | (Sanberg, Morrison and Bjugstad, 2021) | Neural Therapy and Repair | The takeaway message from the 2021 ASNTR conference from society members and attendees was that they missed the in-person experience. | A simple account of the conference | no |
| Craving togetherness: planning and replanning a national society hybrid conference during the COVID-19 pandemic | (Weiniger and Matot, 2021) | Critical care and anaesthesia | 500 attended. “Considering the hybrid format, this compares well with in-person attendee numbers for 2018 and 2019” | Comparison of numbers compared to in person conference. No real analysis | no |
| Hybrid Workshops During the COVID-19 Pandemic-Dawn of a New Era in Neurosurgical Learning Platforms | (Garg et al., 2022) | Neurosurgery | Majority of the respondents (94.1%, n = 64) found hybrid conferences to be better than an online conference. | Not very clear if any people attended online. Only surveyed in person attendees.. | no |
| Review of the Third Conference of the Imaging Mass Spectrometry Society (IMSS 3): Accounts of a Hybrid Virtual and In-Person Meeting and the State and Future of the Field | (Chandler et al., 2022) | Imaging | Post conference survey suggested many attendees were satisfied with the hybrid format but preferred in-person. | They thought that hybrid wouldn’t work for >200 attendees. No analysis of participants and/or questions asked | no |
| Hybrid Conferences in the Post-COVID-19 Era: Time Yet for a Paradigm Shift for Medical Associations | (Devaraj et al., 2022) | Dermatology | Post conference survey found that 82% rated the hybrid conference interaction to be more satisfying than an online conference and 92% found it a safer option. | Simple survey of preference | no |
| "Hybrid" scientific conference: lessons learned from the digital annual meeting of the CARS international conference during the Covid-19 pandemic | (Ostler et al., 2022) | Computer Assisted Radiology & Surgery | Based on the questionnaire, 60% of responders considered the hybrid approach as superior and 12% as inferior to purely virtual conferences. | More a reflection of the logistics of the conference, with a post conference survey added on | no |

**Supplement S6. Characteristics of questions asked by panel composition.** Data presented as n (%).

| PANEL COMPOSITION | MAJORITY FEMALE | MAJORITY MALE | ALL MALE | P | MAJORITY WHITE | ALL WHITE | P |
|---|---|---|---|---|---|---|---|
| n | 77 | 48 | 5 | | 82 | 48 | |
| *Question type* | | | | <0.001 | | | 0.60 |
| Neutral | 37 (48.1) | 30 (62.5) | 1 (20.0) | | 46 (56.1) | 22 (45.8) | |
| Polite | 14 (18.2) | 4 (8.3) | 0 (0.0) | | 10 (12.2) | 8 (16.7) | |
| Shows under-confidence | 3 (3.9) | 3 (6.2) | 0 (0.0) | | 3 (3.7) | 3 (6.2) | |
| Shows over-confidence | 20 (26.0) | 11 (22.9) | 0 (0.0) | | 20 (24.4) | 11 (22.9) | |
| Unknown | 3 (3.9) | 0 (0.0) | 4 (80.0) | | 3 (3.7) | 4 (8.3) | |
| *Mode* | | | | 0.02 | | | 0.04 |
| Microphone | 2 (2.6) | 5 (10.4) | 0 (0.0) | | 2 (2.4) | 5 (10.4) | |
| Chat | 45 (58.4) | 33 (68.8) | 1 (20.0) | | 47 (57.3) | 32 (66.7) | |
| Chair/panel member | 30 (39.0) | 10 (20.8) | 4 (80.0) | | 33 (40.2) | 11 (22.9) | |
| *Gender* | | | | 0.54 | | | 0.36 |
| Female | 44 (57.1) | 30 (62.5) | 4 (80.0) | | 48 (58.5) | 30 (62.5) | |
| Male | 33 (42.9) | 17 (35.4) | 1 (20.0) | | 34 (41.5) | 17 (35.4) | |
| Unknown | 0 (0.0) | 1 (2.1) | 0 (0.0) | | 0 (0.0) | 1 (2.1) | |
| *Observed ethnicity* | | | | 0.59 | | | 0.14 |
| White | 75 (97.4) | 43 (89.6) | 5 (100.0) | | 79 (96.3) | 44 (91.7) | |
| Black | 0 (0.0) | 1 (2.1) | 0 (0.0) | | 0 (0.0) | 1 (2.1) | |
| Asian | 1 (1.3) | 1 (2.1) | 0 (0.0) | | 2 (2.4) | 0 (0.0) | |
| Unknown | 1 (1.3) | 3 (6.2) | 0 (0.0) | | 1 (1.2) | 3 (6.2) | |
| *Country* | | | | 0.34 | | | 0.09 |
| UK | 26 (33.8) | 22 (45.8) | 0 (0.0) | | 24 (29.3) | 24 (50.0) | |
| Europe | 44 (57.1) | 22 (45.8) | 4 (80.0) | | 49 (59.8) | 21 (43.8) | |
| Outside EU | 4 (5.2) | 2 (4.2) | 1 (20.0) | | 6 (7.3) | 1 (2.1) | |
| Unknown | 3 (3.9) | 2 (4.2) | 0 (0.0) | | 3 (3.7) | 2 (4.2) | |
| *Seniority* | | | | <0.001 | | | 0.01 |
| Professor | 28 (36.4) | 7 (14.6) | 2 (40.0) | | 29 (35.4) | 8 (16.7) | |
| Doctor / Senior academic | 34 (44.2) | 31 (64.6) | 0 (0.0) | | 38 (46.3) | 27 (56.2) | |
| Scientist / AHP / PhD student | 3 (3.9) | 0 (0.0) | 2 (40.0) | | 1 (1.2) | 4 (8.3) | |
| Non-medical professional | 9 (11.7) | 4 (8.3) | 1 (20.0) | | 11 (13.4) | 3 (6.2) | |
| Unknown / other | 3 (3.9) | 6 (12.5) | 0 (0.0) | | 3 (3.7) | 6 (12.5) | |

**Supplement S7. Relationship between panel composition by gender and audience participation.** Total 12 sessions: 1 all male, 7 majority (50% or more) female, 4 majority male.

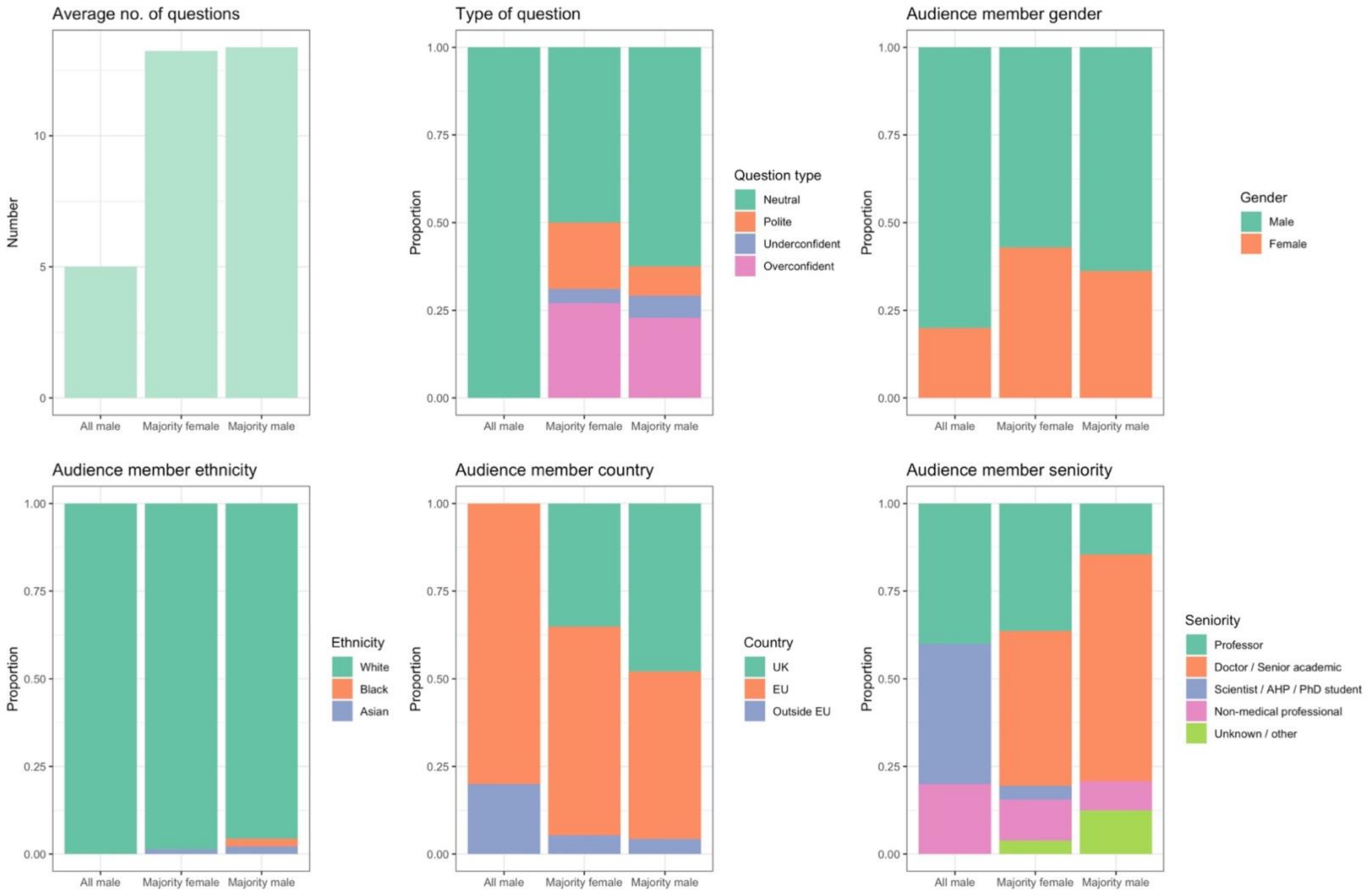

**Supplement S8. Relationship between panel composition by gender and audience participation.** Total 12 sessions: 1 all male, 11 mixed.

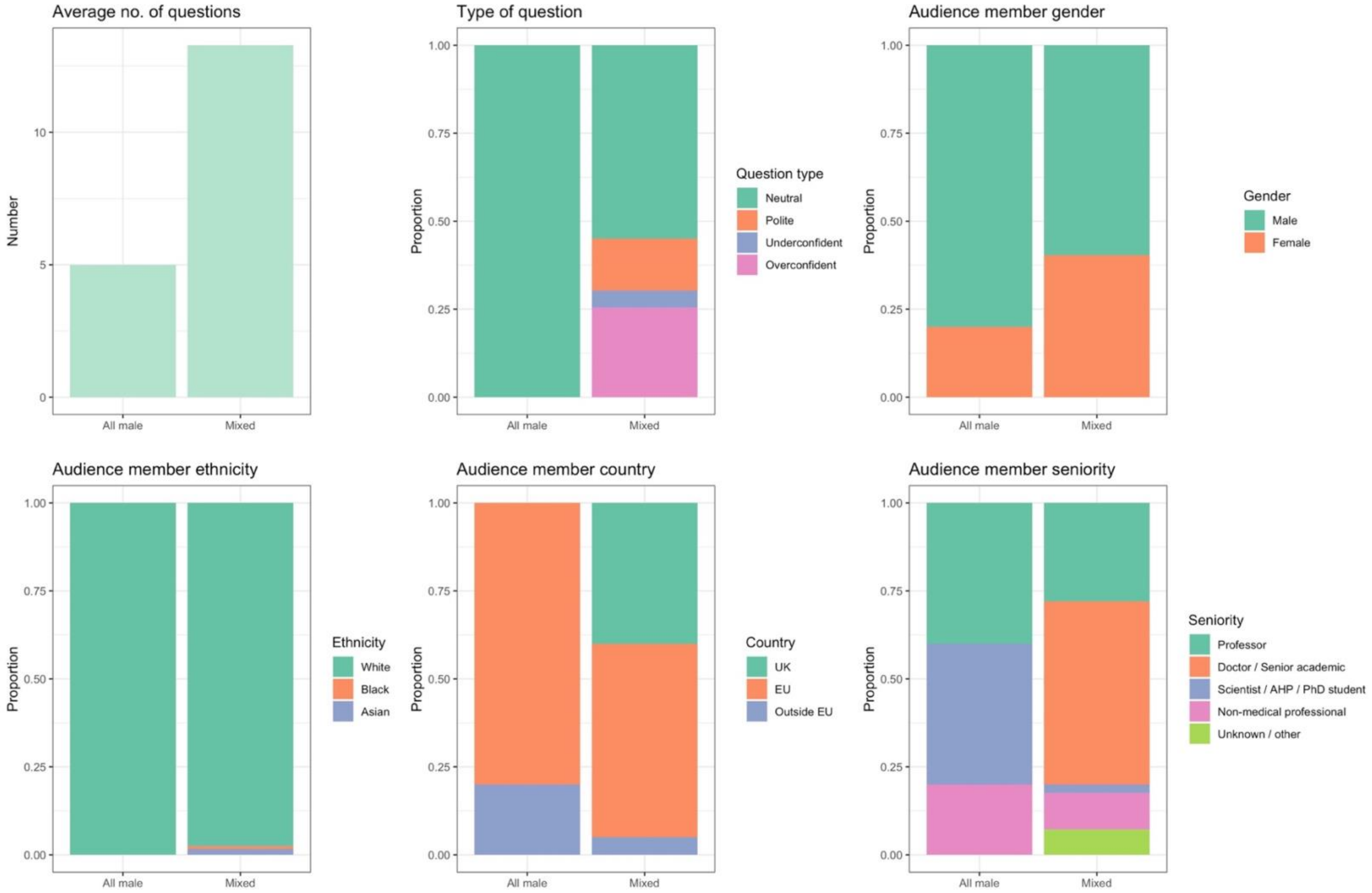

**Supplement S9. Relationship between panel composition by ethnicity and audience participation.** Total 12 sessions: 6 all white, 6 majority white.

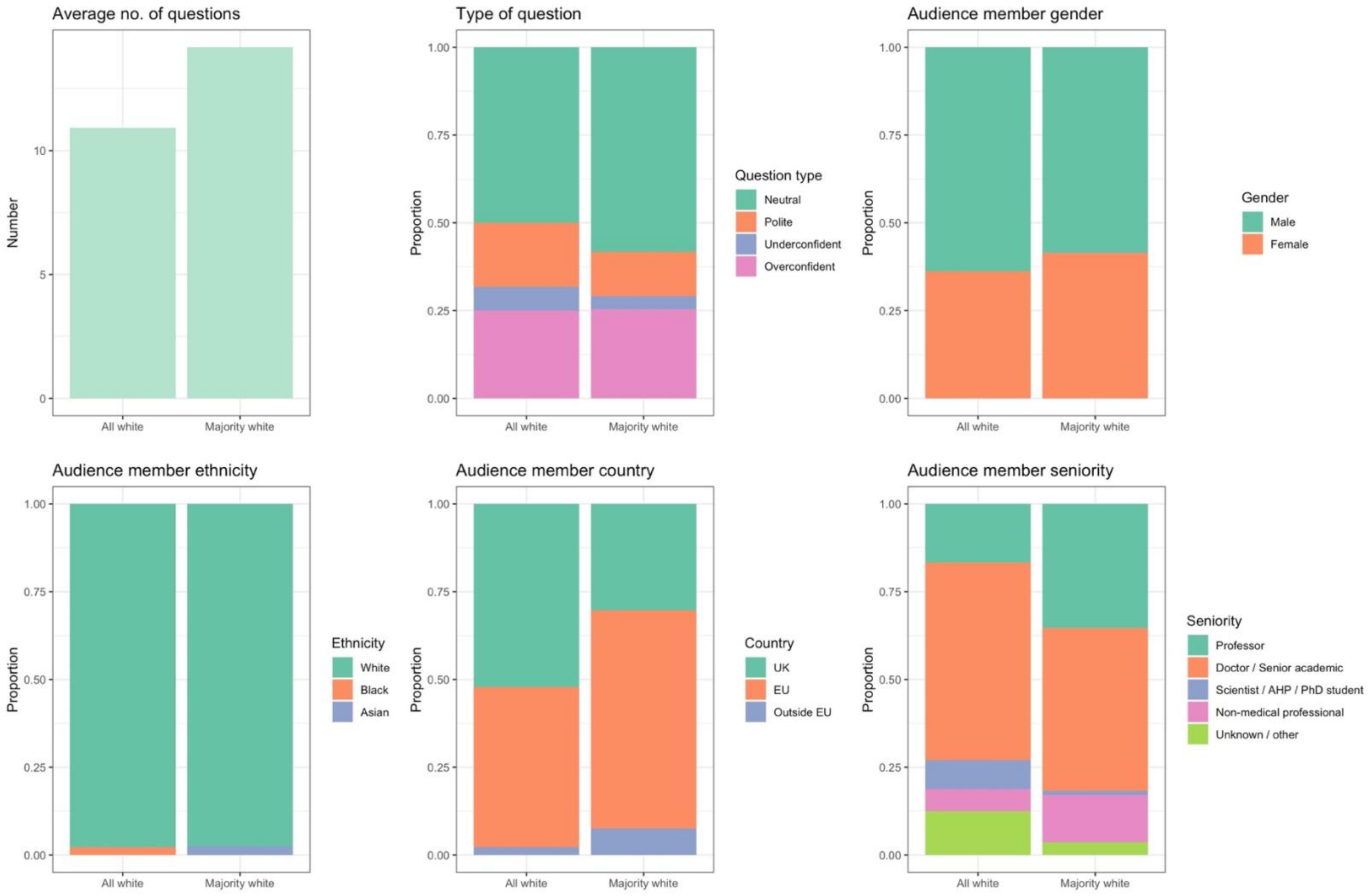

**Supplement S10. Relationship between panel composition by gender and ethnicity and audience participation.** Total 12 sessions: 1 all male all white, 2 majority female all white, 5 majority female majority white, 3 majority male all white, 1 majority male majority white.

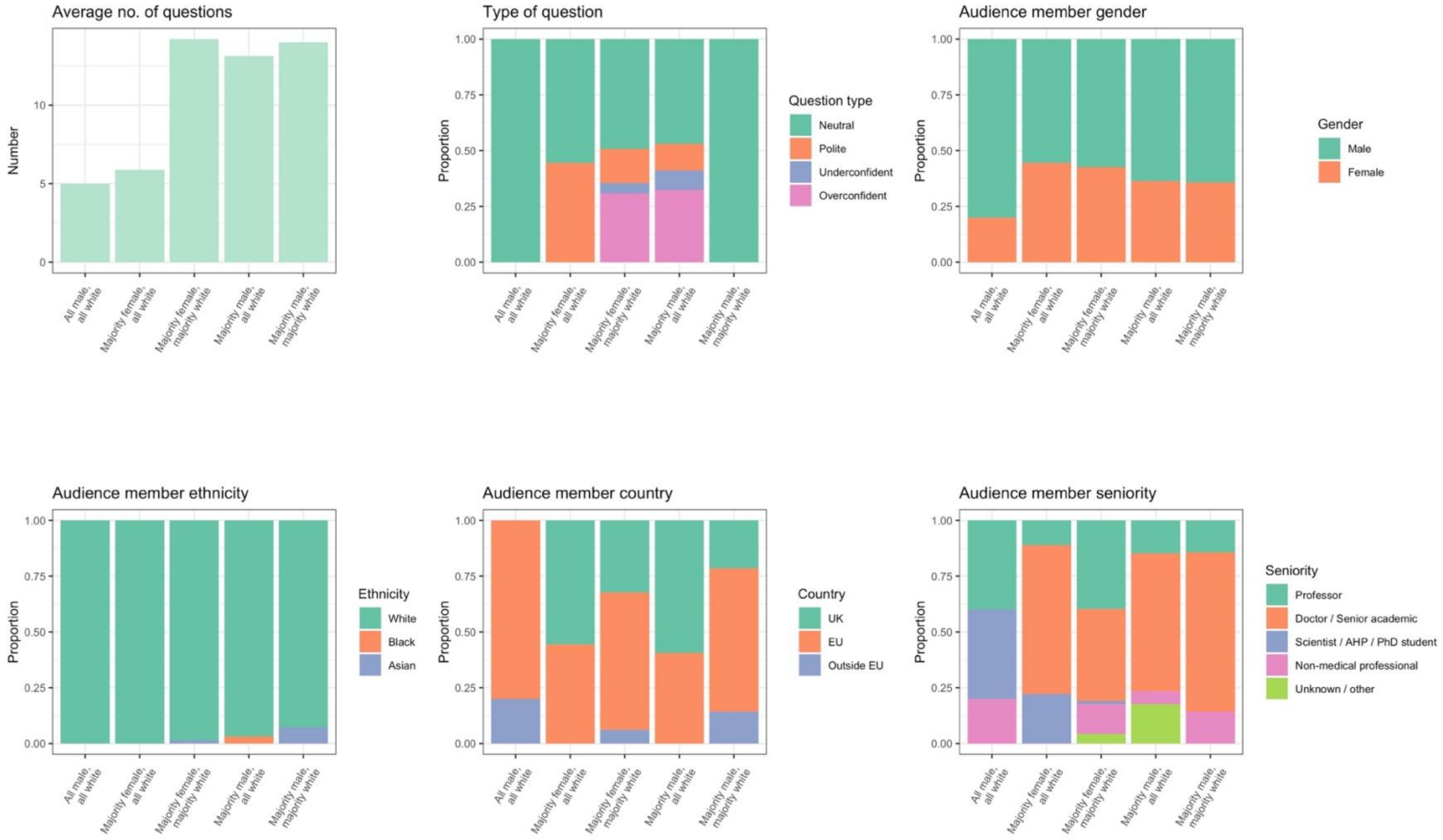
